# Extremely Small Body Size, Not Biological Sex, as a Mortality Risk Factor in Coronary Artery Bypass Grafting: A Nationwide Study in Japan

**DOI:** 10.1101/2025.09.02.25334966

**Authors:** Katsuhiko Oda, Hiraku Kumamaru, Aya Saito, Makoto Takahashi, Shintaro Katahira, Naoko Kinukawa, Yuichi Ueda, Noboru Motomura

## Abstract

**Background:** Coronary artery disease is a significant health concern and can lead to death. Coronary artery bypass grafting (CABG) is the primary surgical treatment for this disease. However, the long-standing hypothesis that women face higher surgical mortality than men after CABG remains controversial. The universal healthcare system and the established national cardiovascular surgery registry of Japan provide a unique oppotunity to assess this supposition. This study aimed to re-evaluate this long-standing hypothesis.

**Methods:** This nationwide observational retrospective study analyzed 40,796 primary elective CABG procedures performed in Japan between January 2019 and December 2023. It included 33,202 men (81%) and 7,594 women (19%). Data were sourced from the Japan Cardiovascular Surgery Database. Preoperative, intraoperative, and postoperative variables were analyzed to assess sex differences in operative mortality. The impact of the yearly procedure volume at each facility was evaluated.

**Results:** Operative mortality for elective CABG was 1.25% in men and 1.63% in women (*P*=0.01), supporting higher operative mortality in women in Japan. Preoperatively, women had lower body surface area and smoking rates. No notable sex differences were observed in the choice of surgical procedure, cardiopulmonary bypass use, or graft selection. Postoperative mediastinitis occurred more frequently in women than in men. After adjusting for body surface area, the difference in mortality became insignificant. A multivariable logistic regression analysis controlling for age, body mass index, preoperative comorbidities, preoperative status, and the facility’s annual CABG procedure volume confirmed that a body surface area of <1.4 m^2^ was a significant mortality risk factor, irrespective of sex. The higher proportion of women with a body surface area of <1.4 m^2^ explained the higher mortality in this group compared to the cases in men. These findings support that extremely small body size is a mortality risk factor for CABG.

**Conclusions:** Nationwide registry data from Japan revealed that female sex is not an independent predictor for CABG mortality. Higher mortality in women primarily reflects the overrepresentation of female patients with very small body sizes. These findings suggest that surgical procedures and anastomosis strategies for small coronary arteries can improve outcomes in all patients when optimized.

## Clinical Perspective

1. What is new?

- This study aims to resolve the long-standing debate on whether women face higher surgical mortality than men after coronary artery bypass grafting (CABG) based on robust national-level evidence, using the Japan Cardiovascular Surgery Database.
- The study revealed that female sex is not an independent predictor for CABG mortality.
- Extremely small body size (body surface area <1.4 m^2^) was a significant mortality risk factor, irrespective of sex.
2. What are the clinical implications?

- In Japan, the unparalleled healthcare infrastructure eliminates key confounding factors, such as sex-based differences in access to care or procedural selection.
- This study represents a globally unique opportunity to re-evaluate the long-standing belief that female sex is a risk factor for CABG mortality.
- The findings of this study suggest that surgical procedures and anastomosis strategies for small coronary arteries can improve outcomes in all patients when optimized.

## Introduction

Coronary artery disease is a significant health concern and can lead to death. Coronary artery bypass grafting (CABG) is the primary surgical treatment for this disease, which is a procedure to anastomose coronary arteries that are 1–3 mm in diameter. It demands advanced technical expertise from surgeons. It has long been considered an established procedure, and the anastomosis technique itself is rarely debated, with good results worldwide. However, whether biological female sex constitutes an independent risk factor for operative mortality in CABG remains a subject of debate.^1,2^ Research across countries highlights five key determinants of female CABG mortality risk: access to medical care, body size, biological structure and physiological function, choice of surgical procedure, and study population size.

The first critical aspect is access to the medical system. The social conditions that women face and their access to healthcare systems vary considerably from country to country.^3–5^ In some countries, not everyone has access to the medical insurance system, making it particularly difficult for women from low-income families to access healthcare. Limited access to healthcare for women can lead to an increase in the number of surgeries required for more severe medical conditions, such as emergencies or congestive heart failure due to low ejection fraction. Concerns exist that this may lead to disparities in medical care and overall treatment, including surgery and perioperative care. Important socioeconomic factors are often not reflected in nationwide registries. In contrast, Japan has a universal health insurance system without economic, regional, or sex disparities in medical access and treatment selection.^6,7^

The second important variable is the body size.^8,9^ Physical size varies greatly between different races and ethnicities.^10^ Women in Europe and North America generally have a larger physique, while in East Asia, especially Japan, the physique tends to be smaller. Smaller body sizes are associated with smaller coronary arteries. Additionally, women tend to have smaller coronary arteries than men when body size is the same.^11^ CABG is a surgical procedure that involves anastomoses of coronary arteries of various diameters. Surgical outcomes are significantly worse in coronary artery anastomosis of <2 mm, especially 1 mm, compared to those in coronary artery anastomosis of ≥2 mm. Consequently, cardiac surgeons must possess a high skill level for coronary anastomosis.^11^ Smaller body size implies narrower coronary arteries, which in turn places higher technical demands on cardiac surgeons. From this perspective, the facilities with a low annual volume of CABG cases may have lower technical abilities and poor outcomes for CABG in patients with smaller body sizes^12,13^.

The third fundamental aspect is the organ structure and physiological function. Three elements of coronary circulation disorders exist: organic or structural stenosis, functional stenosis, and microcirculatory disorder.^14–16^ Particularly, the latter two are more frequent disorders in women. To stabilize the outcome of the CABG in women, it is critical to keep in mind that these disorders exist in the background. This awareness is also required to accurately perform intraoperative graft flow measurements and provide sufficient postoperative drug therapy based on evidence. Sex differences in CABG mortality could be detected if adequate drug therapy, such as calcium blockers, was not available to control these adverse reactions. The universal healthcare system adequately minimizes the differences in perioperative care between men and women in Japan. In addition, intraoperative bleeding and anemia are more frequent in women than in men; this might be the potential cause of the difference in CABG mortality between the two groups.^17,18^

The fourth notable aspect is the selection of the operative procedures.^19–21^ Insufficient revascularization, such as limited use of the left internal thoracic artery (LITA), may contribute to higher CABG operative mortality in women.^19^ Some have argued that such decisions may reflect more operator bias than actual phenotype differences, as the female sex has not been definitively associated with smaller vessels, more diffuse atherosclerotic disease, or longer time requirements to construct distal anastomosis. However, this view seems questionable because smaller patients have proportionally smaller coronary arteries, including the left anterior descending artery (LAD). These smaller coronary arteries pose well-known technical hurdles. In contrast, off-pump coronary artery bypass (OPCAB) may reduce CABG risk in women;^22^ however, this is controversial because OPCAB may increase the difficulty of anastomosis of small coronary arteries in women with small body size.

The fifth vital aspect is insufficient sample size of target patients for the study. When considering women as a risk factor for CABG surgical mortality, it is important to ensure that the study is sufficiently powered. To minimize the effects of economic, regional, and facility disparities within a country, it would be desirable to use a nationwide database or registry if possible. Despite numerous studies addressing sex differences in CABG outcomes, most reports are constrained by limitations in data completeness, socioeconomic confounders, and inconsistent medical access.^19,23^ In contrast, Japan uniquely combines a universal health insurance system with a mandatory, nationwide surgical registry, the Japan Cardiovascular Surgery Database (JCVSD),^24^ ensuring nearly perfect coverage of all CABG cases across the country. JCVSD collects data on all cardiovascular surgical cases at all cardiovascular surgical facilities in Japan through a web-based collecting system established in 2000.

The unparalleled healthcare infrastructure of Japan eliminates key confounding factors, such as sex-based differences in access to care or procedural selection. It allows for the assessment of true biological differences in surgical outcomes. Thus, this study represents a globally unique opportunity to re-evaluate the long-standing assumption that female sex is a risk factor for CABG mortality. We aim to resolve this debate on the hypothesis based on robust national-level evidence. The findings of this study could help develop pragmatic strategies for improving the treatment of coronary artery disease.

## Methods

The Institutional Review Board of Iwate Prefectural Central Hospital approved this nationwide observational retrospective study (approval number: 1839), which conforms to the principles of the Declaration of Helsinki. The requirement for informed consent from patients was waived by adopting an opt-out method.

### Patients, and data source: Collection and verification

Between January 2019 and December 2023, 57,001 consecutive patients with primary isolated CABG were admitted to all cardiovascular surgery institutes in Japan. The numbers of men and women were 45,744 and 11,257, respectively. Data were collected from all over the country using the web-based collecting system of JCVSD.^24^ Dr. Hiraku Kumamaru and Dr. Naoko Kinukawa analyzed the data. The JCVSD organization’s site visit team regularly ensured data reliability.

### Crude operative mortality analysis

First, we evaluated the operative mortality of CABG by sex in groups of overall, emergent, urgent, and elective cases without any adjustment for patient background. To evaluate surgical mortality in a more homogeneous group, we decided to analyze the group of elective cases (male 30202 (81%) and female 7594 (19%) patients).

### Preoperative, intraoperative, and postoperative variables by sex in elective cases

We presented preoperative status data by sex, including congestive heart failure, New York Heart Association classification III and IV, and low ejection fraction (<30%), which may reflect the difficulty in accessing the medical system due to the high proportion of these variables. Body mass index (BMI) and body surface area (BSA) were presented as body size indicators. Furthermore, some Japanese facilities were considered to be extremely low-volume centers. Thus, we demonstrated the distribution of CABG volume (Figure 1) by center. We also presented intraoperative parameters, including operative time, cardiopulmonary bypass time, OPCAB rate, ITA use rate, and the number of anastomoses and postoperative factors, such as mortality and major morbidities.

**Figure 1.**
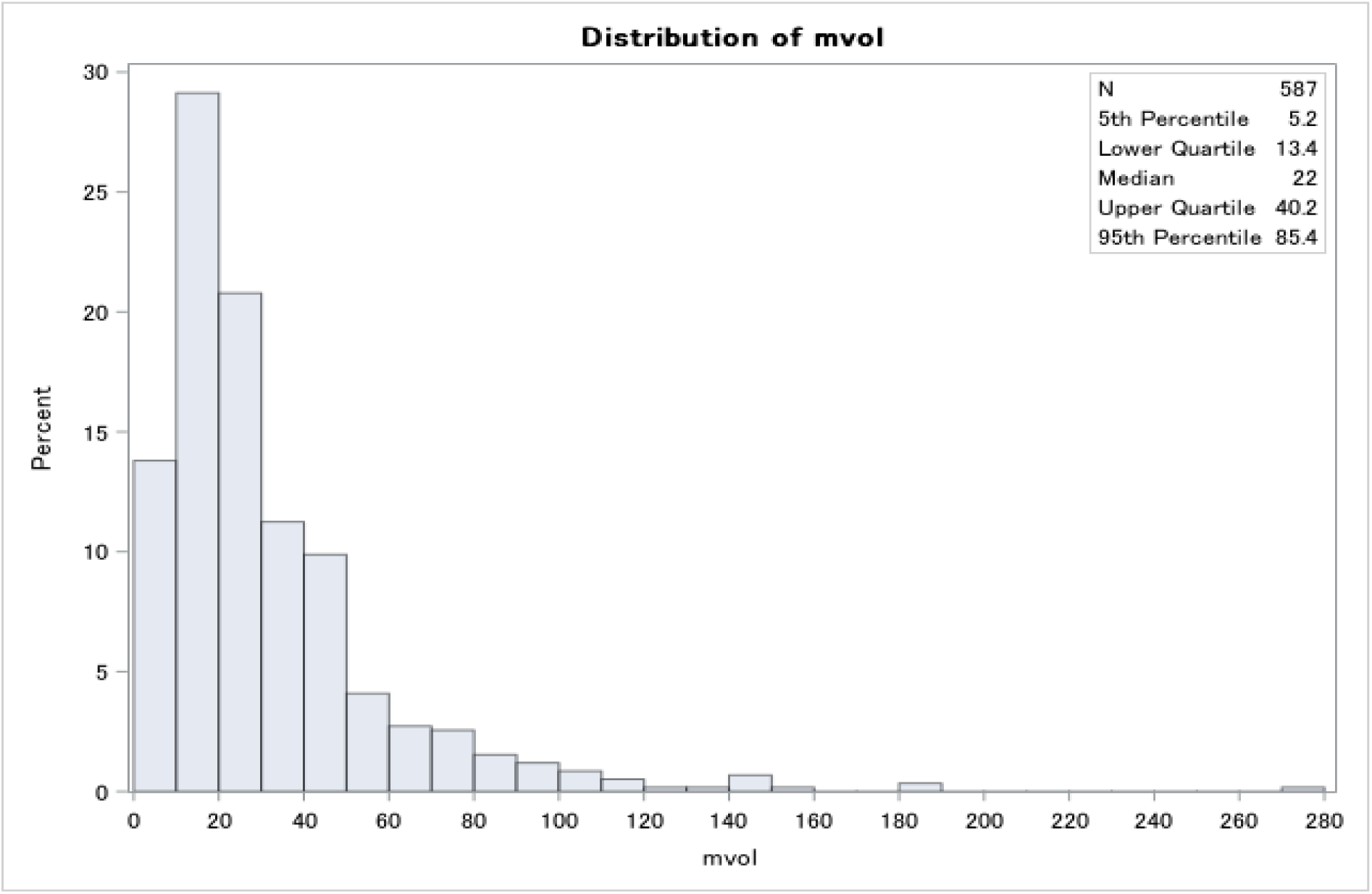
Distribution of facility volume of coronary artery bypass grafting (per year) in Japan.

### Risk factor analysis using multivariable logistic regression

After calculating the odds ratios of operative mortality rates for men and women using crude data in elective cases, we calculated the odds ratios excluding age, BMI, preoperative comorbidities, preoperative status, CABG volume of the facility, and BSA. We searched for items for which the odds ratios regressed to one.

### BSA distribution and outcome analysis

Given the sex difference completely disappeared with BSA in risk factor analysis using multivariable regression, we divided the patients into four BSA groups (BSA ≤1.4, 1.4< BSA ≤1.6, 1.6< BSA ≤1.8, BSA >1.8) and stratified by sex in elective cases. We have shown the distribution and calculated the operative mortality rate for each group.

### Statistical analysis

Continuous variables are presented as medians (interquartile range–(IQR)) and were compared for postoperative maximum creatinine using the Kruskal–Wallis test. Categorical variables are presented as counts and percentages and were compared for operative mortality rate, re-exploration rate for bleeding, rate of neurological complications, rate of mediastinitis, and rate of prolonged ventilation (>72 h) using the Chi-squared test. Multivariable logistic regression was applied for risk factor analysis. The significance level was set at *P*<0.05. All statistical calculations were performed using SAS version 9.4 (SAS Institute, Inc., Cary, NC, USA).

## Results

### Crude operative mortality analysis

Overall, primary isolated CABG operative mortality in women was higher than in men in Japan (3.38% vs. 2.68%, *P*<0.001). Elective CABG operative mortality rate in women was also higher than in men (1.63% vs. 1.25%, *P*=0.01). In contrast, CABG operative mortality rates in women and men were similar in emergent and urgent cases. We adopted a limit for the subsequent analysis of the elective cases.

### Preoperative, intraoperative, and postoperative factors by sex in elective cases

Table 1 shows the preoperative factors: women had lower BSA and smoking rates. Table 2 presents the intraoperative parameters: no notable sex differences were found in the surgical procedure choice, cardiopulmonary bypass use, or graft selection. Table 3 shows the rate of mortality and major morbidities and noted that postoperative mediastinitis occurred more frequently in women.

**Table 1.** Preoperative Patient Characteristics Stratified by Sex Among Patients in Elective Cases.

| Characteristics | Male<br>(n = 33202) | Female<br>(n = 7594) |
| --- | --- | --- |
| Age, year | 70 (62–76) | 74 (68–79) |
| BMI (kg/m<sup>2</sup>) | 23.8 (21.6–26.1) | 23.1 (20.8–25.8) |
| BSA (m<sup>2</sup>) | 1.7 (1.6–1.8) | 1.5 (1.4–1.6) |
| Comorbidities |  |  |
| Diabetes mellitus | 19219 (57.9) | 4447 (58.6) |
| Hypertension | 26366 (79.4) | 6157 (81.8) |
| Glomerular filtration rate (mL/min) | 57.9 (40.6–71.5) | 55.6 (37.6–70.2) |
| Current renal failure requiring dialysis | 3987 (12.0) | 828 (10.9) |
| Smoking history | 19800 (59.6) | 1544 (20.3) |
| Current | 8659 (26.1) | 430 (5.7) |
| Peripheral vascular disease | 6138 (18.5) | 1271 (16.7) |
| Cerebrovascular disease | 2831 (8.5) | 552 (7.3) |
| Severe/moderate chronic lung disease | 1055 (3.2) | 140 (1.8) |
| LV function |  |  |
| Good (EF ≥ 60%) | 13565 (40.9) | 4033 (53.1) |
| Medium (30% ≤ EF < 60%) | 17491 (52.7) | 3266 (43.0) |
| Bad (EF < 30%) | 2146 (6.5) | 295 (3.9) |
| Congestive heart failure | 12539 (37.8) | 3126 (41.2) |
| NYHA functional class |  |  |
| I | 10772 (32.4) | 2199 (29.0) |
| II | 12260 (36.9) | 2970 (39.1) |
| III | 3021 (9.1) | 857 (11.3) |
| IV | 758 (2.3) | 238 (3.1) |
| N/A | 6391 (19.2) | 1330 (17.5) |
| Cardiogenic shock | 102 (0.3) | 46 (0.6) |
| Facility volume of coronary artery bypass cases (per year) |  |  |
| <20 | 5430 (16.4) | 1374 (17.7) |
| 20–39 | 9424 (28.4) | 2110 (27.8) |
| 40–59 | 6956 (21.0) | 1564 (20.6) |
| 60≤ | 11392 (34.3) | 2573 (33.9) |

Values are median (IQR) or n (%) unless otherwise indicated. EF, ejection fraction; LV, left ventricle; NYHA, New York Heart Association; BMI, body mass index; BSA, body surface area; IQR, interquartile tange.

**Table 2.** Surgical Procedures by Sex Among Patients in Elective Cases.

|  | Male | Female |
| --- | --- | --- |
| Operative factors | (n = 33202) | (n = 7594) |
| Operative time (min) | 317 (255–381) | 328 (265–396) |
| Cardiopulmonary bypass use | 12276 (36.97) | 2811 (37.02) |
| Cardiopulmonary bypass time (min) | 142 (112–179) | 146 (114–183) |
| Standard CABG | 6817 (20.5) | 1613 (21.2) |
| Aortic cross-clamp time (min) | 48 (0–105) | 38 (0–108) |
| Beating heart CABG with cardiopulmonary bypass | 5459 (16.4) | 1198 (15.8) |
| Off pump coronary artery bypass | 20926 (63.03) | 4783 (62.98) |
| Graft-related factors |  |  |
| ITA use | 32377 (97.5) | 7296 (96.1) |
| Number of arterial anastomoses | 1 (1–2) | 1 (1–2) |
| Number of saphenous anastomoses | 1 (0–2) | 1 (1–2) |

Values are median (IQR) or n (%) unless otherwise indicated. CABG; coronary artery bypass
grafting; ITA, internal thoracic artery; IQR, interquartile range.

**Table 3.** Mortality and Morbidities Analyzed by Sex Among Patients in Elective Cases.

| Mortality and Morbidities | Male<br>(n = 33202) | Female<br>(n = 7594) | *P* Value |
| --- | --- | --- | --- |
| Overall mortality rate | 414 (1.25) | 124 (1.63) | 0.01<sup>a</sup> |
| Mortality rate (On pump) | 249 (1.7) | 55 (1.6) | 0.87<sup>a</sup> |
| Mortality rate (Off pump) | 165 (0.9) | 69 (1.6) | <0.001<sup>a</sup> |
| Re-exploration for bleeding | 393 (1.18) | 92 (1.21) | 0.84<sup>a</sup> |
| Neurological complications | 456 (1.37) | 102 (1.34) | 0.84<sup>a</sup> |
| Mediastinitis | 251 (0.76) | 112 (1.47) | <0.001<sup>a</sup> |
| Postoperative maximum creatinine | 1.14 (0.92–1.64) | 0.9 (0.7–1.36) | <0.001<sup>b</sup> |
| Prolonged ventilation (>72 hrs) | 451 (1.36) | 118 (1.55) | 0.19<sup>a</sup> |

Values are median (IQR) or n (%) unless otherwise indicated. <sup>a</sup>Chi-squared test. <sup>b</sup>Kruskal–
Wallis test. IQR, interquartile range.

### Risk factor analysis using multivariate regression

The difference in mortality was no longer significant after adjustment for BSA. A multivariable logistic regression analysis controlling for age, BMI, preoperative comorbidities, preoperative status, and the annual CABG procedure volume of the facility confirmed that a BSA of <1.4 m^2^ was a significant mortality risk factor, irrespective of sex (Table 4).

**Tables 4.**
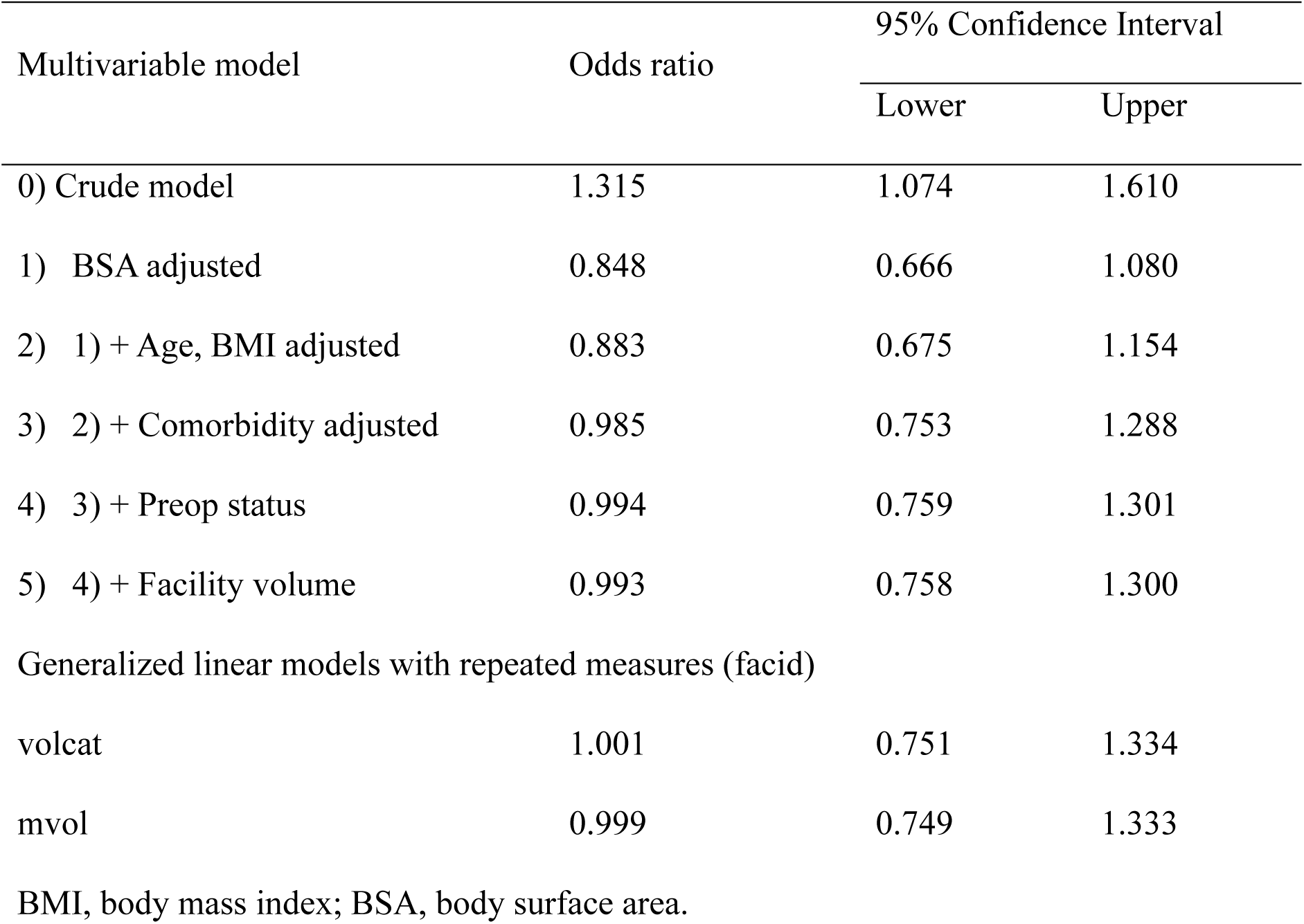
Risk Factor Analysis (Multivariable Model) in Elective Cases.

### BSA distribution and outcome analysis

Figure 2 shows the BSA distribution by sexes of patients who underwent CABG in Japan. Figure 3 shows the relationship between BSA and operative mortality in CABG by sex in Japan. The higher proportion of women with BSA <1.4 m^2^ explained the higher mortality in this group compared to the cases in men. These findings support that extremely small body size is a mortality risk factor for CABG.

**Figure 2.**
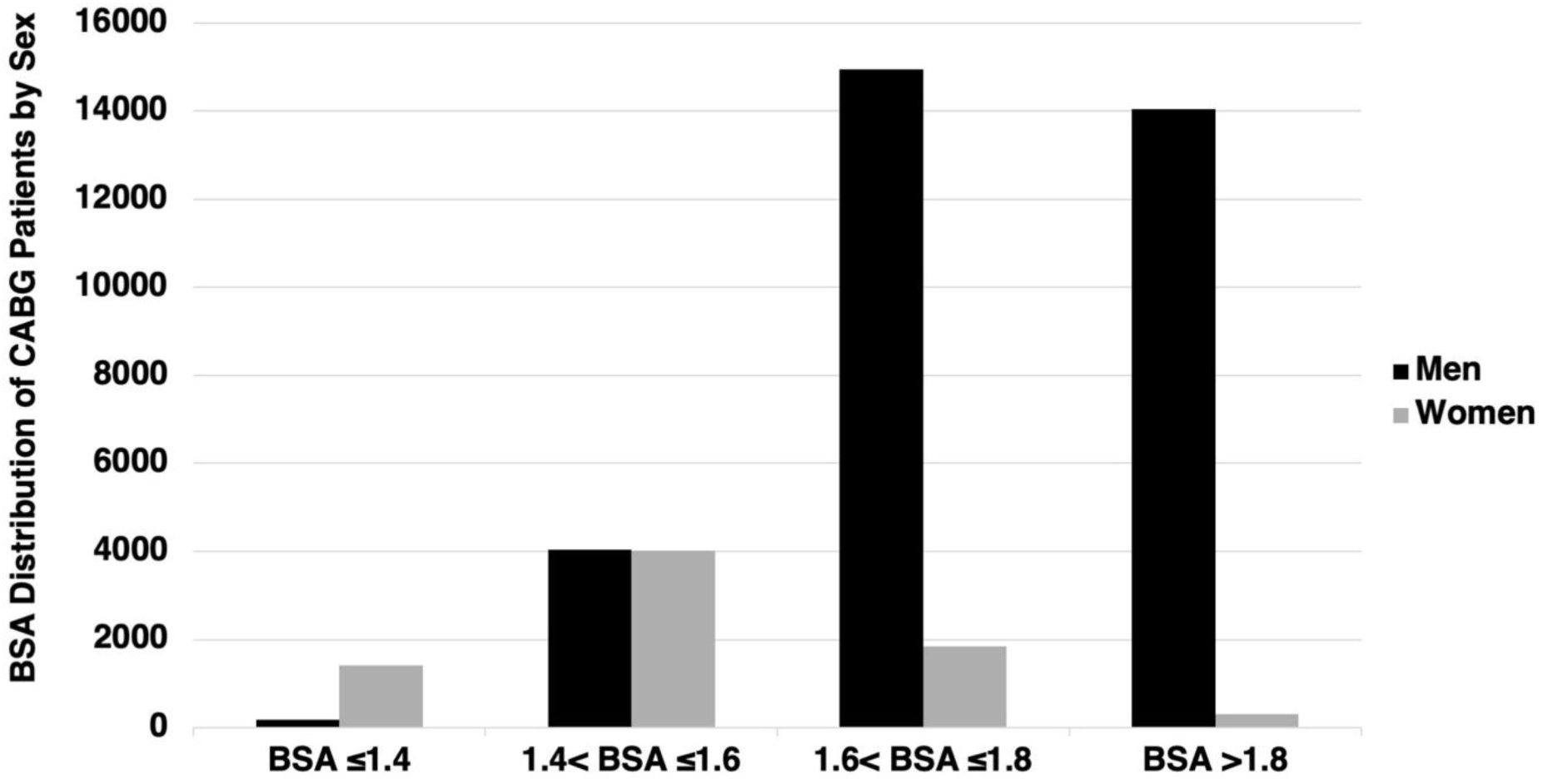
Distribution of body surface area by sex in Japan.

**Figure 3.**
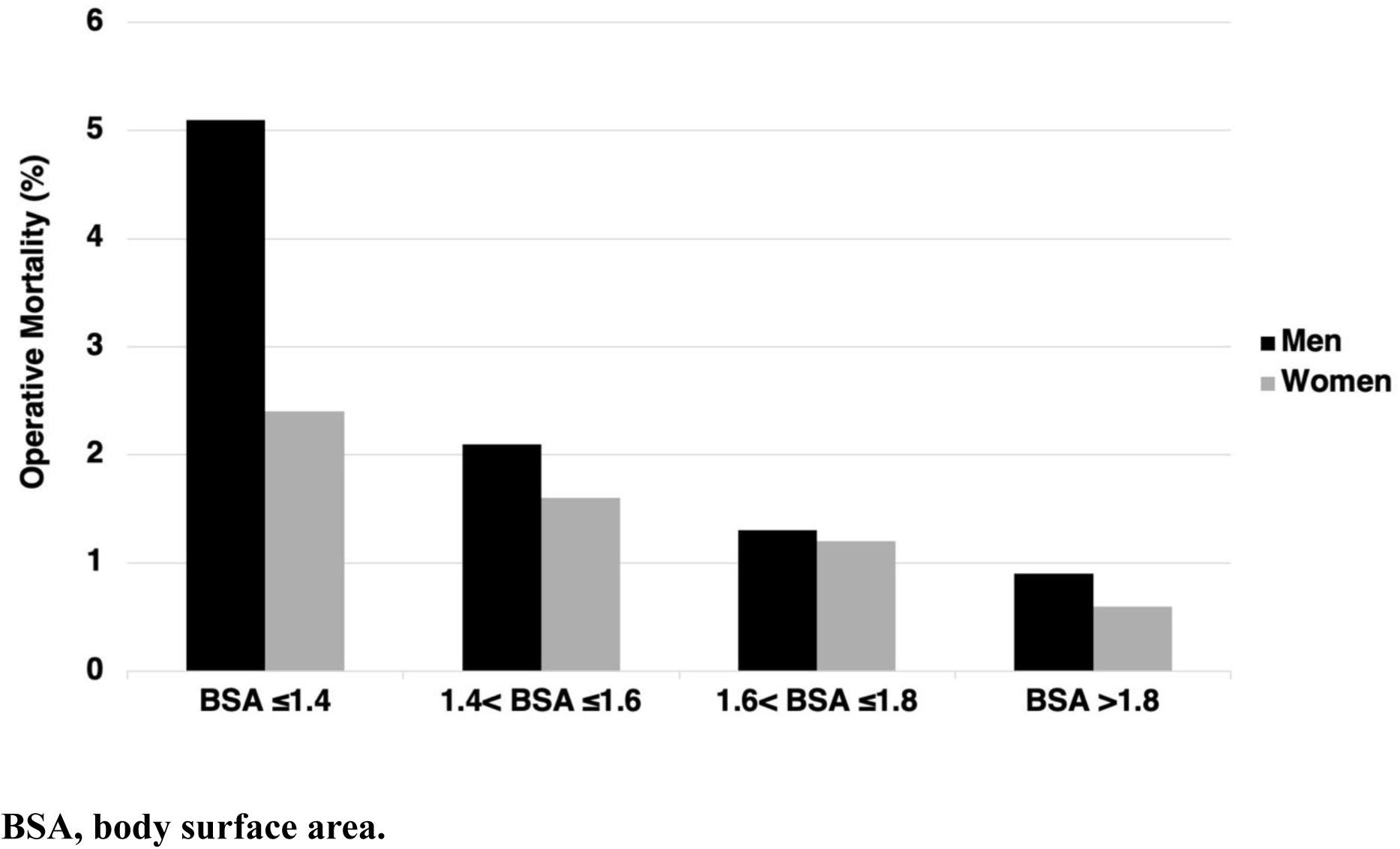
Elective coronary artery bypass grafting mortality versus body surface area.

## Discussion

Nationwide registry data from Japan revealed that female sex is not an independent predictor for CABG mortality. The higher observed mortality in women is largely attributable to the overrepresentation of female patients in the very small body size group. Despite including extremely low BSA, the operative mortality of CABG in Japan was smaller than in other countries, even in OPCAB cases. The Japanese universal health insurance system reduces the differences in access to the medical care and preoperative variables between women and men, excluding BSA and smoking habits. The female-specific physiological dysfunction was not evident during perioperative management, and the choices of surgical procedures was almost the same. Thus, results from the Japanese society and the nationwide JCVSD analysis were suitable to help disqualify the hypothesis that women face higher surgical mortality than men after CABG. It also became clear that the real problem we needed to solve is how to treat small (1 mm) coronary arteries in patients with extremely small body sizes.

Looking into differences due to biological sex differences, the five key items (access to medical care, body size, biological structure and physiological function, choice of surgical procedures, and size of the research individuals) can be divided into three: access to medical care, body size, and size of the research sample. This is because the difference in biological structure and physiological function can be minimized by adequate access to medical care, and the reasons for not using LITA can be explained by the small body size. Therefore, this study, conducted in a country with equitable access to healthcare and a sufficiently large national sample, was particularly suitable for evaluating the impact of body size on CABG outcomes. Considering these three items, we think it is important to evaluate research based on the “sex difference in CABG” of various countries, identify factors contributing to sex differences at various facilities, and make improvements. Conversely, no biological sex difference in CABG mortality exists. Considering body size, medical access, and research scale, identifying the cause of the “apparent sex difference” will lead to the development of appropriate countermeasures.

Pioneers in the 1960s first developed CABG, and in the 1970s, the LITA-LAD + sequential Great Saphenous Vein (GSV) procedure was established using instruments still in use today, such as the Castroviejo needle holder, microscopes, and monofilament polypropylene sutures. The LITA-LAD + sequential GSV procedure achieved good surgical outcomes.^25–30^ Cardiopulmonary bypass,^31^ advances in drug therapy such as calcium channel blockers during the perioperative period,^32^ and developing medical devices used for anastomosis^26,33^ contributed to establishing this stable outcome. The operative mortality for CABG had fallen to less than 1% in the late 1970s.^8^ Furthermore, LITA-LAD have been established as the gold standard for CABG.^34,35^

Initially, anastomosis techniques were actively discussed, particularly in LITA-LAD.^36^ However, for larger coronary arteries (over 1.5 mm) that most patients have, the procedure became straightforward for most cardiac surgeons, leading to a decline in interest in improving the technique. The introduction of percutaneous coronary intervention (PCI) in the 1990s led to significant efforts to enhance CABG techniques. These advancements aimed to improve long-term outcomes while reducing the invasiveness of the procedure. Strategies included using multiple arterial grafts, such as the right internal thoracic, radial, and gastroepiploic arteries, eliminating the need for cardiopulmonary bypass and minimizing the size of surgical incisions.^25,26^ Although these attempts have achieved some success, most cardiac surgeons around the world remain skeptical of them and still perform a LITA-LAD + sequential GSV procedure using a median sternotomy and cardiopulmonary bypass, with good surgical outcomes. Surprisingly, this simple, old-fashioned procedure, which was presumed to be mainly held in the SYNTAX trial, has been shown to have better long-term outcomes in patients with three-vessel disease than PCI.^37^ However, to further improve surgical outcomes, some reports argues that cardiac surgeons should thoroughly follow the guidelines and make extensive use of arterial grafts, particularly in women.^19^

Female sex was recognized as one of the risks for CABG mortality in the 1980s.^38^ However, it gradually became clear that its impact differed depending on socioeconomic context and body size. Some previous reports argued that female sex is not a mortality risk for CABG. Most of these reports come from countries with larger body-sized women (BSA > 1.7 m^2^), including a single institution in the US. ^39–44^ Women in these countries were estimated to have larger coronary arteries (>2 mm), which is less technically complex. However, other reports from the US using the STS database or large registries paradoxically debated that the female sex is an independent mortality risk factor for CABG, although most of women in the US have a large body size.^2,9,19,38,45^

If small body size is an inherent risk factor for CABG mortality, then attributing higher mortality rates to female sex alone may obscure this underlying issue. As a result, necessary countermeasures might be delayed, or inappropriate strategies may be implemented. In countries where the patients with extremely small body sizes are rare, the problem may be overlooked due to analyses showing “no statistically significant difference between men and women.” However, we argue that the impact of small body size should be recognized as a potential global issue. Moreover, in countries with a higher proportion of larger patients, where the hypothesis that female sex independently increases CABG mortality has not been refuted, attention should be directed toward another crucial factor, inequalities in access to medical care, which must not be ignored.

In Japan, the majority of medical institutions perform only 10–20 CABG procedures per year (Figure 1). Despite this, the country has achieved surgical outcomes comparable to those in Europe and the US. CABG in patients with extremely small body size is routinely performed, and coronary artery anastomoses less than 1.5 mm in diameter are by no means rare. One Japanese institution has reported no significant sex differences in CABG mortality, suggesting that successful anastomosis of small coronary arteries is feasible when adequate technical expertise is present.^46^ In a nationwide analysis, the operative mortality for patients with a BSA of <1.4 m^2^ was 5.1% for men and 2.4% for women. This result indicated that most patients survived the procedure, demonstrating that, even in anatomically challenging patients, the majority of procedures are performed safely, highlighting the high technical proficiency of Japanese cardiac surgeons. Nevertheless, there remains room for improvement in outcomes among patients with very small coronary arteries. To further reduce mortality, systematic education and technical certification system in sub-1.5 mm, especially 1 mm, coronary artery anastomosis techniques may be necessary. While Japanese surgeons currently travel to Europe or the US to study CABG, it may now be more effective to focus on mastering advanced techniques domestically, where experience with anatomically demanding cases is abundant.

Figure 4 illustrates the technical differences between small (1 mm) and large (>1.5 mm) coronary artery anastomosis. In the distal anastomosis procedure for the coronary artery, the anastomosis is performed in an end-to-side manner. This involves longitudinally incising the coronary artery, allowing the graft to be slightly turned outward (everted) before attached. The goal is to ensure that the intima—the innermost layer of each vessel—maintains contact with each other during anastomosis. The smaller the bite on the intimal edge of the coronary artery, the larger the resulting opening can be. However, there is a limit to this, as it becomes difficult to control bleeding if the intimal bite is too small. If the inner diameter of the coronary artery is 1 mm (Figure 4A), the circumference is approximately 3 mm. If the intimal bite is 1mm, the portion remaining on the bottom side is 1mm, and the anastomosis will not function. A suitable bite must be less than 0.3 mm, which is technically demanding but achievable for expert surgeons. In contrast, if the inner diameter is 1.5 mm (Figure 4B), the circumference is approximately 4.5 mm. Performing the anastomosis is straightforward if the intimal bite is 1 mm and the remaining portion on the bottom side is 2.5 mm. Hence, saving patients with small body size requires a skill level that allows easy execution of a 1 mm coronary anastomosis.

**Figure 4.**
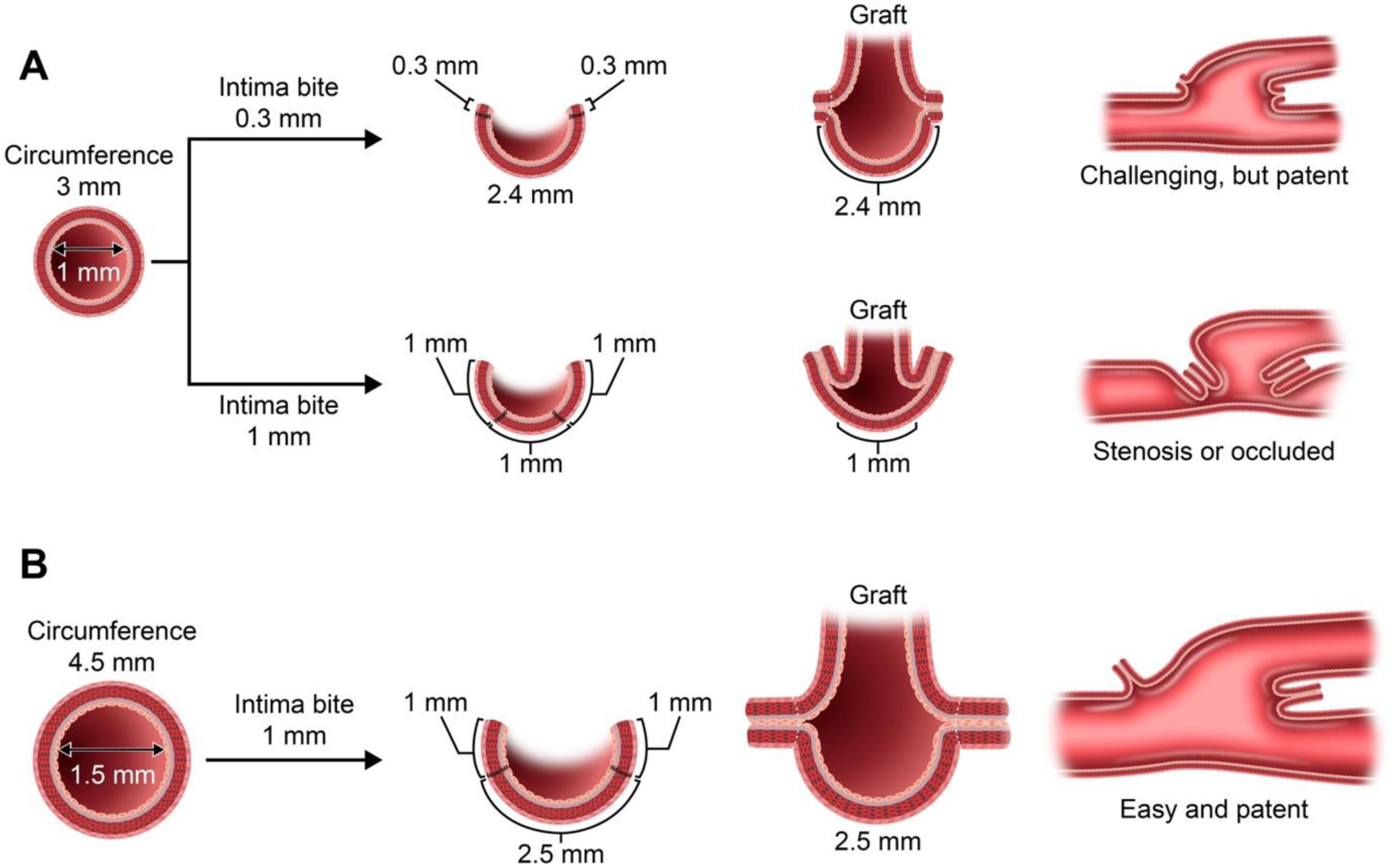
Technical aspect of coronary artery anastomosis: 1 mm versus 1.5 mm.

Another strategy to save patients with small body sizes is to improve the quality of the LITA-LAD. In some cases, it may be beneficial not to anastomose the smaller (<1 mm) lateral, posterior, or inferior wall branches. LAD may be the largest and most important artery for improving myocardium ischemia because it can perfuse the anterior wall. The diagonal and septal branches that originate from LAD can perfuse the lateral and inferior walls of the left ventricle as collateral sources. If the LAD is 1 mm in diameter, the other branches are often smaller (<1 mm). These branches may be too thin to ensure adequate flow; therefore, avoiding anastomosis and preserving their flow is better. High-grade anastomosis of LITA-LAD can perfuse the entire wall of the left ventricle via collateral flow and rescue patients; however, it may cause “incomplete revascularization.” The high-grade LITA-LAD indicates that thick LITA is anastomosed with the more proximal LAD. LITA should be harvested until it intersects below the left innominate vein without intercostal branches. If bilateral ITAs are not available because of thinness (<1 mm), GSV can be used for LAD without hesitation. This high-grade LITA-LAD strategy is suitable for patients with extremely low BSA.

The quality control of anastomoses and graft preparation is important for CABG in patients with extremely low BSA. In Japan, transit time flowmetry is commonly used to assess graft performance. If coronary surgeons detect low mean graft flow, high pulsatility index, and low diastolic filling, they should perform re-anastomosis without hesitation.^47^ Graft preparation is also important. In the past, papaverine hydrochloride, a potent vasodilator, was used; however, this drug is acidic and poses a risk of endothelial damage. As a result, neutral vasodilators, such as nitroglycerin, calcium channel blockers, and phosphodiesterase 3 inhibitors, are now commonly used.^48^

CABG had already matured as a procedure by the 1970s. However, during the prolonged controversy with PCI, culminating in the final report of the SYNTAX trial in 2019, the true essence of what should have been pursued was lost. Rather than focusing on the technical challenges of small coronary anastomosis, efforts were directed toward increasing the use of arterial grafts, eliminating cardiopulmonary bypass, and minimizing incisions. Now that the superiority of CABG for complex coronary lesions has been reaffirmed in the post-SYNTAX era, this study offers a renewed direction for where our efforts should be concentrated. Essential issues that were overlooked in the evolution of cardiac surgery must be brought back into focus, namely, further enhancement of the quality of LITA-LAD anastomosis (which lies at the core of CABG’s superiority), the establishment of reliable techniques for small coronary arteries, and a strategic reconsideration of whether secure revascularization of the LAD should take precedence over complete revascularization. We hope that this study will reignite attention to these forgotten priorities and ultimately contribute to saving more lives through truly patient-centered, technically sound coronary surgery.

This nationwide, retrospective observational study has some limitations. First, the JCVSD lacks detailed anatomic and functional data, which may have resulted in unmeasured confounding sex-related outcomes. Second, as with all large-scale registries, the risk of reporting bias and data entry errors cannot be excluded. Finally, the impact of the coronavirus disease 2019 (COVID-19) pandemic on cardiac surgery outcomes, particularly during the year 2020, should be considered as a potential confounding factor when interpreting the results.

In summary, nationwide registry data from Japan revealed that female sex is not an independent predictor for CABG mortality. The higher observed mortality in women is primarily due to the overrepresentation of patients with very small body sizes. These results suggest that optimizing surgical procedures and anastomosis strategies for small coronary arteries improves outcomes in all patients.

## Data Availability

All data are not available because the data access of Japan Cardiovascular Surgery Database is strictly limited.

## Acknowledgments

We thank Editage (www.editage.com) for the English language editing.

## Sources of Funding: **None**

**Disclosures:** None

## Non-standard Abbreviations and Acronyms

BMI: body mass index
BSA: body surface area
CABG: coronary artery bypass grafting
GSV: great saphenous vein
IQR: interquartile range
ITA: internal thoracic artery
JCVSD: Japan Cardiovascular Surgery Database
LAD: left anterior descending artery
LITA: left internal thoracic artery
OPCAB: off-pump coronary artery bypass
PCI: percutaneous coronary intervention
STS: The Society of Thoracic Surgeons

## Notes

### Competing Interest Statement

The authors have declared no competing interest.

### Clinical Trial

This is a nationwide, retrospective observational study, not a clinical trial or a prospective interventional study.

